# Race and gender disparity of dietary lycopene intake and periodontitis severity in older adults

**DOI:** 10.1101/2024.05.29.24308111

**Authors:** Katherine Kwong, You Lu, ZhuoHuan Li, Ting Luo, Zhaoyu Huang, Na Zhao, Tung-Sung Tseng

## Abstract

**Purpose:** Periodontitis in older adults is a public health issue. There is a growing population of older adults in the United States; furthermore, the proportion of the population of older adults who identify as a racial minority are growing at a faster rate than non-minority identifying older adults. Sufficient lycopene intake is one potential solution for individuals who express reluctance and/or an inability to access preventative oral care particularly by older adults who self-identify as a racial minority, leading to lower levels of periodontitis if they consume a sufficient amount of lycopene. The objective of this paper is to explore the association between insufficient lycopene intake and risk of periodontitis among older adults.

**Methods:** Data analysis was conducted using data from the National Health and Nutritional Examination Survey 2009-2014. Overall, 1227 adults aged 65-to 79-years-old were investigated. The total lycopene intake from daily diet, age, race/ethnicity, living condition, smoking status, body mass index, diabetes, gender, and education level were studied as independent variables. The combination of probing depth and number of interproximal sites was used as criteria for evaluating the risk of periodontitis. Weighted multinominal logistic regression was used.

**Results:** Overall, 48.7% of older adults in this study were classified with different levels of periodontitis and 49.1% of individuals with a form of periodontitis had insufficient dietary lycopene intake.Sufficient lycopene intake was found to be associated with a lower likelihood of severe periodontitis (OR: .33; 95% CI: 0.17-.65; p=0.002). Non-Hispanic Black older adults were more likely to develop severe forms of periodontal disease in comparison to Non-Hispanic White older adults (OR: 2.82, 95% CI: 1.46-5.45, p=0.003). Gender was also found to play a role in periodontitis status, with women being less likely to have severe periodontitis status, compared to men (OR: 0.27; 95% CI: 0.14-0.55; p= 0.0007,). However, only Non-Hispanic White females were less likely to have severe periodontitis compared to Non-Hispanic White males (OR: 0.26; 95% CI: 0.12-0.56; p= 0.001).

**Conclusion:** Dietary intake of lycopene associated with periodontitis disease for individuals over the age of 65; those who consume a sufficient amount of lycopene are less likely to develop severe periodontitis. In general, more men suffered from periodontitis in comparison to women. Such association also has been found between Non-Hispanic Blacks with severe periodontitis. In addition, reduced risk of severe periodontitis is associated with sufficient lycopene intake is explored in Non-Hispanic Whites, future targeted interventions using lycopene in dietary intake as a preventative measure to delay or prevent the onset of periodontal disease needs to be race and gender specific.

## Introduction

It is estimated that periodontal disease affects more than 47% of adults over the age 30 in the United States but affects a significantly higher proportion of older adults over the age of 65, approximately 70% [1]. Periodontitis (PD) As one type of periodontal disease, periodontitis (PD) is characterized by severe inflammation of the gums and if left untreated, can lead to tooth loss. Preventative measures in developing PD include regular oral care from a profession [2-4].While some individuals may be more genetically predisposed to develop PD, PD has the potential to affect any individual with poor dental hygiene [5, 6]. The consequence of tooth loss or edentulism due to untreated PD has the potential to subsequently negatively impact an individual’s nutritional intake [3, 7]. In addition, the loss of a tooth or teeth has also been shown to negatively impact self-esteem and overall quality of life of individuals [8, 9]. Previous studies have observed racial disparity of PD in non-Hispanic White (NHW) and non-Hispanic Black (NHB) adults with an indication that PD is a risk factor for other chronic, and potentially more serious health conditions resulting in more than just edentulism [10-13]. Poor periodontal health is associated with an increased prevalence of other conditions such as: hypertension, diabetes, and stroke [10-12].

While predisposing variables such as age, gender, and race/ethnicity impact observed disparity in PD, they cannot be modified. However, dietary intake of lycopene, a non-provitamin A carotenoid, is one potential preventative measure that can be modified to prevent the development of PD. Tomato products are the primary dietary source of lycopene in the U.S. but it can be found in other fruits or vegetables that have a red or pink hue such as apricots, grapes, guava, and papaya [14, 15]. Extensive studies have established a significant correlation between lycopene as an adjunct to professional dental cleanings and PD [16-18]. To date, there is limited research on how the intersections of race and gender affect dietary consumption of lycopene of minority older adults with severe PD. Understanding the prevalence of PD in NHB older adults and the impact that consumption of lycopene has on PD will inform future strategies for targeted interventions. Further research is needed to not only determine if consumption of lycopene through either nutritional sources or as a supplement is statistically significant in preventing PD and subsequently other conditions associated with PD, but also how dietary lycopene consumption of different groups are impacted in different ways.

## Methods

We combined and extracted three consecutive two-year survey cycle datasets (2009-2014) from the U.S. Centers for Disease Control NHANES dataset for this study. A total of 1227 adults aged between 65 and 79 years-old, with completed response to questions on lycopene intake and a recorded oral health record, were included. The lycopene consumption level from dietary intake was collected from a two-day dietary interview questionnaire on total nutrient intakes, based on recall responses from survey participants. A multistage, stratified, probability-cluster sampling method was used during the NHANES data collection, under the supervision of the National Center for Health Statistics of the CDC [19]. An in-person, face-to-face interview was conducted for qualified participants at their homes by trained staff. The oral health examination was conducted in a mobile examination center (MEC). Individual demographic and health-related information was collected via examination. The survey protocol was revised and approved by the National Center for Health Statistics Research Ethics Review Board [20]. A paper-based informed consent form was signed by every participant before the data collection process was initiated.

### Measurements

The main exposure of interest for this work was lycopene intake. NHANES uses the five-step U.S. Department of Agriculture’s Automated Multiple Pass Method (AMPM) to collect dietary intakes, including lycopene. More details on AMPM are provided in the NHANES dietary interviewer procedure manuals. A clinical study has shown that lycopene is a promising treatment for patients with moderate periodontal disease [16]. We defined sufficient lycopene intake from daily food as >=8000mcg [21]. Anyone who consumes lycopene but <8000 mcg was defined as lycopene intake insufficient. Three levels of periodontitis are defined as follows [22]: severe was defined as having >=2 interproximal sites with attachment loss (AL) >= 6 mm (not on the sample tooth) and >=1 interproximal sites with probing depth (PD) >= 5mm; non-severe periodontitis was defined as combination of moderate and mild cases such that: as >=2 interproximal sites with AL >=4 mm (not on the same tooth), or >=2 interproximal sites with PD >=5mm or one site with PD >=5mm (moderate); or >=2 interproximal sites with AL >= 3 mm and >= 2 interproximal sites with PD >= 4mm (not the same tooth) or one site with >=5 mm (mild); none was defined as meets neither the severe nor non-severe case definitions. Smoking status was categorized into never smoker, former smoker, and current smoker based on the following questions: “Have you smoked at least 100 cigarettes?” and “Do you now smoke cigarettes?”. Never smoker was defined as a respondent who reported that they smoked <100 cigarettes in their lifetime; former smoker was defined as respondents who reported smoking >= 100 cigarettes in their lifetime and currently do no smoke cigarettes; current smoker was defined as respondents who reported smoking >=100 cigarettes in their entire lifetime and were currently smoking every day or some days [23]. The demographic information of included participants included age (65∼69, 70∼79), race/ethnicity (NHW, NHB), gender, and education (less than high school or more than high school). Individuals without diabetes are defined as diabetes free. Based on the martial status options, a participant who selected “married” or “living with a partner” was defined as living with a partner; a participant who selected “widowed”, “divorced”, “separated”, or “never married were defined as living alone [24]. In addition, Body-Mass-Index (BMI) was classified as underweight/normal, overweight and obese, respectively [25].

### Ethics

The procedures to obtain human data have been performed in accordance with the Declaration of Helsinki. All methods carried out in this study for the purpose of this analysis were reviewed and approved by the Institutional Review Board of Connecticut College #2023.24.23.

### Outcomes

The level of periodontitis was the primary outcome of interest for this study. We used a combination of probing depth and attachment loss to determine periodontitis status.

### Statistical Analysis

Participants’ demographics, lycopene intake, smoking behavior, and clinical features were summarized using descriptive statistics based on the different PD statuses (severe to none). The association of these factors with PD status was tested using the Rao-Scott Chi-square test for categorical variables and Fisher’s exact test for small samples. A one-way ANOVA test was applied to examine differences for continuous variables. All analyses were performed in R (svydesign) using the “survey” package [26] to create the weighted analysis groups for displaying percentages (%) (svyciprop) and means with standard error of the mean (svymean). Survey-weighted multinominal logistic regression models (svymultinom) were applied to examine the association between lycopene intake and PD status in older adults by using the “nnet” and “svrepmisc” package. We included the suggested covariables: age, gender, race, smoking status, and education to construct the multinominal logistic regression model [27-34]. All variables had no multicollinearity in the model. We followed the weighting instructions provided on the CDC website [35]. All analyses were conducted using R version 3.6.3 with the packages that mentioned above and repeated in Python 3.8 with package “samplics” [36]. All tests were 2-sided, and a P-value <0.05 was considered statistically significant for all tests.

## Results

A total of 1227 NHW and NHB older adults between the ages of 65 and 79 were included from NHANES 2009-2014. Table 1 provides weighted percentage and raw sample sizes from demographics and lycopene intake by level of PD. For the overall study population, 22.1% of participants (n=246) had sufficient lycopene intake; 77.9% of participants (n=981) had insufficient lycopene intake. The self-reported daily lycopene intake of participants with severe, non-severe, and no level of PD is 3847±360, 5452±498, 5278±338 (mcg), respectively (p=.006). The prevalence of all levels of periodontitis is 48.7%. NHB older adults have a higher prevalence of severe PD and overall PD in comparison to NHW older adults (12.2% vs 4.86% and 55.6% vs 47.86%, respectively; p=.0004). The mean age of people diagnosed with severe PD is 69.9±0.4, compared to other groups (p=0.17). Although NHB only takes 10.5% of the whole participants, around 12.2% of them have been diagnosed with severe PD, which is almost three-fold more than NHW (p=0.0004). Older adults who take sufficient lycopene from daily dietary have less people diagnosis with severe PD (2.4% vs 6.5%, respectively, p=0.04). The severe PD ratio is also high in current smokers (18.8%), in which is 4-fold more compared to never and ever smoker (4.6%, 4.1%, respectively, p=0.0001). In addition, less old female adults have been found in severe PD, compared to older male adults (3.1% vs 8.3%, respectively, p=0.001). Overall, lycopene intake, gender, smoking, and race are risk factors that significantly contribute to the level of PD in this study.

**Table 1.** Factors associated with different degrees of periodontitis in older adults aged 65 years and older (Weighted)

| Characteristics | Overall | PD Sev | PD Non Sev | Normal | p-Value |
| --- | --- | --- | --- | --- | --- |
| total, n(%) | 1227 | 98(5.6) | 531(43.1) | 598 (51.3) |  |
| Mean lycopene Intake ± SEM (mcg) | 5273(318) | 3847(360) | 5452(498) | 5278(338) | 0.006 |
| Mean age ± SEM (year) | 70.5(0.1) | 69.9(0.4) | 70.6(0.2) | 70.5(0.2) | 0.17 |
| Lycopene Intake, n(%) |  |  |  |  | 0.04 |
| Insufficient | 981(77.9) | 81(6.5) | 424(42.6) | 476(50.9) |  |
| Sufficient | 246(22.1) | 17(2.4) | 107(44.7) | 122(52.9) |  |
| Race, n(%) |  |  |  |  | 0.0004 |
| Non-Hispanic White | 875(89.5) | 51(4.86) | 377(43) | 447(52.1) |  |
| Non-Hispanic Black | 352(10.5) | 47(12.2) | 154(43.4) | 151(44.4) |  |
| Education Level, n(%) |  |  |  |  | 0.14 |
| < High School | 269(16) | 25(5.5) | 98(35.6) | 146(58.9) |  |
| ≥High School | 956(84) | 73(5.7) | 431(44.3) | 452(50.0) | |
| Smoking Status, n(%) |  |  |  |  | 0.0001 |
| Never Smoker | 553(47.1) | 41(4.6) | 233(43.8) | 279(51.6) |  |
| Ever Smoker | 542(44.2) | 38(4.1) | 253(44.5) | 251(51.3) |  |
| Current Smoker | 120(8.7) | 19(18.8) | 42(34.9) | 59(46.3) |  |
| Gender, n(%) |  |  |  |  | 0.001 |
| Male | 624(48.1) | 70(8.3) | 288(45.3) | 266(46.3) |  |
| Female | 603(51.9) | 28(3.1) | 243(40.9) | 332(56.0) |  |
| Age , n(%) |  |  |  |  | 0.73 |
| 65-70 | 570(52.9) | 51(6.1) | 257(43.9) | 262(50.0) |  |
| 71-79 | 657(47.1) | 47(5.1) | 274(42.1) | 336(52.8) |  |
| Living Condition, n(%) |  |  |  |  | 0.53 |
| With Partner | 769(69.4) | 57(5.5) | 330(42.2) | 382(52.4) |  |
| Alone | 458(30.6) | 41(6.0) | 201(45.1) | 216(48,9) |  |
| Diabetes, n(%) |  |  |  |  | 0.89 |
| Diabetes Positive | 288(18.2) | 19(6.9) | 129(45.6) | 140(47.5) |  |
| Diabetes Negative | 886(77.2) | 75(5.1) | 380(42.7) | 431(52.2) |  |
| BMI, n(%) |  |  |  |  | 0.79 |
| Under/normal Weight | 340(29.5) | 31(4.6) | 144(43.8) | 165(48.8) |  |
| Overweight | 457(37.2) | 36(4.8) | 200(43.9) | 221(51.2) |  |
| Obese | 423(33.3) | 31(6.5) | 42(40.0) | 59(53.5) |  |

The computed results from weighted multinomial logistic regression models are given in Table 2. After adjusting for covariables, participants with sufficient lycopene intake have 0.33 time odds (95% CI: 0.17-0.65, p=0.002) associated with severe PD, compared to old adults who have insufficient lycopene intake. NHB has 2.82 times odds (95% CI: 1.46-5.45, p=0.003) associated with severe PD in comparison to NHW. Female participants have 0.27-time odds of severe PD, compared to male participants (95% CI: 0.14-0.55, p=0.0007). Further moderation analysis confirmed race as a moderator between dietary lycopene intake and severe PD (p< 0.0001). Results of stratification analysis between NHW and NHB are given in Table 3. For NHW, female participants have 0.26-time odds (95% CI: 0.12-0.56, p=0.001) associated with severe PD compared to males. And NHW participants with sufficient lycopene intake has 0.13-time odds associated with severe PD (95% CI: 0.05-0.37, p<0.0001), compared to the insufficient intake one. Such association between server PD and gender/lycopene intake did not observe in NHB.

**Table 2.** Factors associated with PD status at severe (Sev), non-severe (PD Non Sev) to non-PD.

| Characteristics | PD Sev |  |  | PD Non Sev |  |  |
| --- | --- | --- | --- | --- | --- | --- |
|  | Adjusted OR | 95%CI | p-Value | Adjusted OR | 95%CI | p-Value |
| Lycopene Intake |  |  | 0.002 |  |  | 0.9 |
| Insufficient (Ref) | 1 |  |  | 1 |  |  |
| Sufficient | 0.33 | 0.17-0.65 |  | 0.97 | 0.64-1.49 |  |
| Race |  |  |  |  |  |  |
| Non-Hispanic White (Ref) | 1 |  | 0.003 | 1 |  | 0.121 |
| Non-Hispanic Black | 2.82 | 1.46-5.45 |  | 1.32 | 0.93-1.87 |  |
| Smoking Status |  |  |  |  |  |  |
| Never Smoker (Ref) | 1 |  |  | 1 |  |  |
| Ever Smoker | 0.7 | 0.33-1.48 | 0.33 | 0.99 | 0.63-1.54 | 0.95 |
| Current Smoker | 3.29 | 1.55-6.97 | 0.002 | 0.86 | 0.44-1.70 | 0.66 |
| Education Level |  |  | 0.4 |  |  | 0.09 |
| < High School (Ref) | 1 |  |  | 1 |  |  |
| >= High School | 1.49 | 0.58-3.84 |  | 1.5 | 0.93-2.42 |  |
| Gender |  |  | 0.0007 |  |  | 0.09 |
| Male (Ref) | 1 |  |  | 1 |  |  |
| Female | 0.27 | 0.14-0.55 |  | 0.73 | 0.50-1.06 |  |
| Age |  |  |  |  |  |  |
| 65-70(Ref) | 1 |  |  | 1 |  |  |
| 71-79 | 0.91 | 0.46-1.82 | 0.78 | 0.9 | 0.57-1.43 | 0.65 |

**Table 3.** Factors associated with PD across different race groups.

|  | Non-Hispanic Black |  |  |  |  |  |
| --- | --- | --- | --- | --- | --- | --- |
| Characteristics | PD Sev |  |  | PD Non Sev |  |  |
|  | Adjusted OR | 95%CI | p-Value | Adjusted OR | 95%CI | p-Value |
| Lycopene Intake |  |  | 0.15 |  |  | 0.41 |
| Insufficient (Ref) | 1 |  |  | 1 |  |  |
| Sufficient | 2 | 0.76-5.26 |  | 0.73 | 0.34-1.57 |  |
| Gender |  |  | 0.13 |  |  | 0.87 |
| Male (Ref) | 1 |  |  | 1 |  |  |
| Female | 0.37 | 0.11-1.37 |  | 0.94 | 0.44-2.03 |  |
|  | Non-Hispanic White |  |  |  |  |  |
| Characteristics | PD Sev |  |  | PD Non Sev |  |  |
|  | Adjusted OR | 95%CI | p-Value | Adjusted OR | 95%CI | p-Value |
| Lycopene Intake |  |  | <0.0001 |  |  | 0.95 |
| Insufficient (Ref) | 1 |  |  | 1 |  |  |
| Sufficient | 0.13 | 0.05-0.37 |  | 1..01 | 0.69-1.54 |  |
| Gender |  |  | 0.001 |  |  | 0.08 |
| Male (Ref) | 1 |  |  | 1 |  |  |
| Female | 0.26 | 0.12-0.56 |  | 0.72 | 0.51-1.03 |  |

## Discussion

For the overall study population, the majority consumed an insufficient amount of dietary lycopene (77.9%). However, in addition to dietary lycopene consumption, other risk factors of severe PD were: race, gender and smoking status with NHB, men and current smokers being at higher risk of developing severe PD. The results of this analysis continue to add to the literature that there is variability between NHB and NHW older adults with regard to PD (https://aap.onlinelibrary.wiley.com/doi/full/10.1002/JPER.22-0137). NHB women had higher odds of severe PD in comparison to NHW and race was confirmed to be a moderator between lycopene intake and severe PD. Previous research has suggested several biological and social factors that may be contributing to this observed racial disparity. Genetic predisposition to PD, the inflammation and acid production of the body after the consumption of sugary foods that can potentially lead to gum disease. Aggressive forms of PD have been shown to have familial aggregation within the NHB community which suggests that these individuals may be genetically predisposed to develop PD [37]. The results of this analysis continue to add to the literature that there is variability between NHB and NHW older adults with regard to PD [13]. NHB women had higher odds of severe PD in comparison to NHW and race was confirmed to be a moderator between lycopene intake and severe PD. Previous research has suggested several biological and social factors that may be contributing to this observed racial disparity. Genetic predisposition to PD, the inflammation and acid production of the body after the consumption of sugary foods that can potentially lead to gum disease. Aggressive forms of PD have been shown to have familial aggregation within the NHB community which suggests that these individuals may be genetically predisposed to develop PD [38].

However, the conclusion of this analysis indicates that another potential factor explaining the racial disparity in the development of PD is that there is an observed disparity in dietary lycopene intake. Lycopene intake is significantly higher in NHW, while prevalence of severe PD is lower [39]. Another possible reason why NHB older adults had higher prevalence of PD may be due to how additional preventative measures of PD were determined for the purpose of this analysis. Previous studies recognize racial differences in oral health status, but few emphasize the disparity of PD in older adults and analyze modifiable risk factors that focus on nutrition instead of preventative dental care or non-modifiable factors [40-42].

The present study also investigated whether gender impacted the prevalence of PD. The likelihood of having PD was significantly lower for older females than males. Females were found to be approximately half as likely to develop severe PD non-severe PD than males (Odds Ratio = 0.27 and 0.73). Men have been found to be less likely to seek preventative dental care, use tobacco products at a higher rate, and hormone differences [43]. Additionally, race mediated the impact of dietary lycopene intake and PD. Higher levels of lycopene consumption were associated with a lower likelihood of PD in older adults. However, this study has determined that sufficient lycopene intake is associated with a higher odds ratio of severe PD. Potential reasons for this discovery could potentially be the known irreversibility of PD. Once PD is diagnosed, individuals can work to prevent the increased severity of the disease that may lead to eventual tooth loss but they cannot reverse the current progress of PD. Subsequently, individuals who have been informed that they have mild PD and are at risk for developing severe PD, may act to reduce their risk of developing a more severe form of the disease.

A comprehensive review of the literature conducted by Leite (2018) concludes what many studies have indicated, that smoking has a detrimental effect on PD and the progression of PD [44]. The disparity in prevalence of smoking amongst different race/ethnicity groups in the US is well documented [45, 46]. However, dietary lycopene intake has been shown to be a protective agent against the effects of tobacco promoted changes such as the development of non-alcoholic steatohepatitis in animal models [47]. Subsequently, although the prevalence of smoking in this study population is 8.7%, dietary lycopene intake is a potential factor in the reason why there remains an observed racial and gender disparity in observed severe PD.

There are several limitations to this analysis. First, although PD is easily preventable with regular professional oral care, and dental insurance has the ability to alleviate some of the costs associated with preventative treatment, the NHANES dataset does not record the status of dental insurance [48, 49]. Health insurance status, which is recorded by NHANES, cannot be used as a stand in for dental insurance because preventative dental insurance is not necessarily included in general health insurance. Although all individuals included in this analysis are over the age of 65 and therefore eligible for Medicare coverage, Medicare does not cover dental services in most cases, services such as routine dental cleanings [50]. Older adults also have the opportunity to choose Marketplace coverage via the Affordable Care Act (ACA) instead of Medicare if they have to pay a Part A premium. However, the ACA treats dental coverage differently based on the age of the enrolled individual, where dental coverage is an essential benefit for individuals under the age of 18 and must be available, it is not an essential benefit for older adults and health plans on the Marketplace are not required to offer older adults dental coverage [51]. This is one of the challenges encountered when conducting a secondary data analysis with the NHANES dataset, one of the best preventative measures, preventative oral care, is unable to be considered in this study because the data was not collected. Although previous studies have shown that preventative dental treatments have lowered the risk of developing PD, this factor was not included in this study. There are numerous external factors that other studies have linked to the prevention of PD that were not considered for the models of analysis in this study that may have an additional impact in not only the prevalence of PD, but also an individual’s consumption of dietary lycopene.

Although lycopene intake was only calculated as total dietary intake and the source of lycopene was not taken into account the conclusion of this study is that there is a relationship between prevalence of severe PD and lycopene intake, which potentially contributes to the observe racial/ethnic and gender disparity between NHW and NHB older adults and PD. However, this analysis cannot make the determination if there is a difference between lycopene consumption and PD from food sources versus a combination of lycopene consumption from food and supplements. The distinction between intake of lycopene from a food source in contrast to lycopene from a supplement, may have an impact on PD due to the fact that previous studies have demonstrated that antioxidant treatment with lycopene could potentially be beneficial as an adjunctive treatment periodontal disease [52]. Nevertheless, certain foods that contain lycopene may actually impact inflammation and acid production in the oral cavity which may have an impact on the prevalence of gum disease due to either increased levels of lycopene in the blood or due to increased levels of lycopene in the oral cavity. While this study concludes that there is a correlation between dietary intake of lycopene and severe PD in older adults, the vehicle in which lycopene impacts PD in this analysis is not determined.

## Conclusion

The objective of this study is to explore the association between insufficient lycopene intake and the risk of PD in older adults. Overall, insufficient lycopene intake was found to be a risk factor associated with the development of PD. Although this study was not able to distinguish between the different sources of dietary lycopene, the conclusions of this analysis determined that dietary intake of lycopene is a predictor of severe PD for NHW individuals between the ages of 65 and 79 years old. Subsequently, although dietary lycopene intake was found to be associated with levels of PD in this analysis, the racial differences indicate that while Future targeted interventions using lycopene in dietary intake as a preventative measure to delay or prevent the onset of periodontal disease needs to be race and gender specific.

## Data Availability

All data produced are available online at https://www.cdc.gov/nchs/nhanes/index.htm

## Author Contributions

K.K. and Y.L. contributed equally to this study. K.K. contributed to the original literature review and to the conception and design of this study. Y.L. contributed to the conception, design, and database organization of the study. Y.L. and ZH.L. performed the statistical analysis in R. ZY.H performed the statistical analysis in Python. K.K. and T.L cross-validated this study. T.S.T. and N.Z. supervised this study. All authors contributed to manuscript writing.

## Data and code Availability

Publicly available datasets were analyzed in this study. These data can be found at: https://www.cdc.gov/nchs/nhanes/index.htm. The analysis code will be provided upon request.

## Ethics approval

The study used publicly available datasets that meet the federal regulation at 46 CFR 46.102.

## Consent to participate

N.A.

## Consent to publish

N.A,

